# The role of Body Mass Index in high and low velocity trauma causing knee injury associated to popliteal artery lesions

**DOI:** 10.1101/2019.12.31.19015966

**Authors:** A Ascoli Marchetti, V Naldi, V Potenza, F.M. Oddi, F. Di Maio, G. Citoni, P Farsetti, A. Ippoliti

**Affiliations:** Orthopedic and Traumatology Unit, University of Rome Tor Vergata, Surgical Sciences Department

## Abstract

**Background:** Osteoarticular traumas are particularly dangerous among arterial traumas, those associated with the popliteal artery are associated with a high amputation rate. Despite representing a minority of arterial traumas, with an incidence that considerably varies by population and geographic location, traumatic lesions of the popliteal artery are a challenge. This study aimed to verify the impact of BMI on arterial trauma damage and outcome.

**Methods:** Data were retrospectively collected from the emergency and operating rooms’ electronic medical reports at our Institution between 1 January 2005 and 1 May 2018 of all osteoarticular and vascular associated lesion. 41 pts presented with lower limb arterial trauma (43.2%), and popliteal artery lesions occurred in 11 (26.8%). 11 patients were eligible for inclusion in the study. In addition, the lesion mechanism was dislocation by high-velocity trauma in 9 cases and by low-velocity trauma in 3. All 7 males (63.6%) were affected by high-velocity trauma, and 2 of the 3 females by low-velocity trauma. Only one patient had an isolated popliteal artery lesion associated with fractures in the leg or in contralateral limb. Patients with low-velocity traumas were older than 54 years while those with high-velocity were aged from 22 to 71 years.

**Results:** The lesion mechanism was dislocation due to high-velocity trauma in eight patients and due to low-velocity trauma in three. In 10/11 patients (90.9%) revascularization was performed after osteoarticular stabilization. after reduction of the dislocation or fracture. Intraoperative angiography was selectively used. Two patients required above-knee amputation after the procedure: one due to infection of the surgical access and the other due to severe soft tissue injury. One patient died during hospitalization due to trauma-related complications and comorbidities.

**Conclusions:** Revascularization success is not associated with high- or low-velocity traumas. Furthermore, unlike high-velocity traumas, low-velocity traumas are associated with a body mass index >35kg/m2.

## Background

Traumatic lesions of the popliteal artery are rare1,2, with an incidence rate between 5% and 19% in the civilian population3,4. Because of its anatomical position in the popliteal fossa, because of which it is anteriorly protected by the knee joint, it is barely affected by isolated traumas. It is more frequently associated with knee fractures (Gustillo IIIC)5,6 or knee dislocation7,8. Ligation of arterial injuries of the leg in World War II led to an amputation rate of 72%9, and experience with arterial repair or reconstruction in the Korean War lowered the amputation rate to 32%10. More than a decade later, similar amputation rates were reported from the Vietnam War11. Although a much lower amputation rate has been reported in years between 1990 and 2000, traumatic lesions of the popliteal artery continue to be the arterial lesion most associated with limb loss12,13. A multidisciplinary approach is still key in the successful management of knee injury. Dislocation or fracture can cause a vascular lesion and must be recognized quickly for imminent repair. The osteoarticular cause must also be identified for appropriate revascularization. Surgeons should always consider the possibility of a popliteal artery lesion, even in cases of minor trauma14,15. Few studies focus on the relationship between speed and mass. This study aimed to evaluate arterial injury-associated risk factors of trauma and verify the role of body mass on the outcome of surgical arterial revascularization.

## Methods

Prior to initiating the writing of this document, all patients and/or family members were contacted to seek their consent for the release and processing of sensitive data for research purposes. Main inclusion criteria were traumatic lesions of the popliteal artery and a patient older than 18 years at the time of admission. Exclusion criteria was arterial lesion not involving limbs. Another criterion was absence of a diagnosed popliteal artery lesion (such as popliteal artery aneurysm) prior to trauma and admission to the hospital. The patients were divided into two study groups based on the dynamics that produced the arterial lesion: “high” or “low” velocity. A high-velocity injury was one in which the trauma involved a motorized vehicle, such as motor vehicle collisions, motorcycle collisions, moped collisions, motorized vehicle collisions, or crush injuries. A low-velocity injury was defined as trauma occurring secondary to a fall, a sport, or an assault8,14. Clinical and operative data were retrospectively collected from the emergency and operating rooms’ electronic medical reports at Tor Vergata Hospital between 1 January 2005 and 1 May 2018. Ninety five patients were admitted to Tor Vergata Hospital with a diagnosis of arterial trauma (Table 1). Inclusion criteria was popliteal artery injury, so of these, 42 presented with lower limb arterial trauma (44.2%) and in these cases popliteal artery lesions occurred in 11 (26.8%). 10 patients were eligible for inclusion in the study. In addition, the lesion mechanism was dislocation by high-velocity trauma in 9 cases and by low-velocity trauma in 2. All 7 males (70%) were affected by high-velocity trauma, and 2 of the 3 females by low-velocity trauma. Three patients had popliteal artery lesions and knee dislocation (all posterior dislocation), two of whom had a total knee prosthesis; five patients had knee fractures associated with popliteal artery trauma and one patient had knee fracture and dislocation involving the popliteal artery. Only one patient had an isolated popliteal artery lesion associated with fractures in the leg or in contralateral limb. Patients with low-velocity traumas were older than 54 years while those with high-velocity were aged from 22 to 71 years.

**Table 1.** Arterial injuries admitted to our institution betwenn 2005 and 2018.

| *Injury mechanism* | **Overall group** | **CFA lesion** | **DFA lesion** | **SFA lesion** | **Popliteal artery lesion** | **Tibial arteries lesion** |
| --- | --- | --- | --- | --- | --- | --- |
| *Number* | 42 | 8 | 5 | 13 | 11 | 5 |
| *Gunshot wound* | 3 | - | - | 3 | 1 | - |
| *Motorized vehicle accident* | 21 | 4 | 4 | 6 | 6 | 1 |
| *Stab/laceration* | 6 | 1 | 1 | 3 | - | - |
| *Crush injury* | 7 | 3 | - | 1 | 1 | 2 |
| *Fall* | 4 | - | - | - | 3 | 2 |
| *Assault* | - | - | - | - | - | - |
| *Sport* | - | - | - | - | - | - |

## Results

All patients were treated within 3 hours from admission to the emergency room and within 6 hours from the accident. In eight patients, revascularization was performed after osteoarticular stabilization, except for one patient in whom revascularization was performed before orthopaedic surgery due to bleeding from the artery. One patient only required arterial reconstruction without subsequent collaboration with orthopaedists due to the absence of fractures or dislocations. In most part of the popliteal area involvement, 9/11 (81,8%) were used posterior surgical access to the knee, with only two patients (18.1%) requiring the medial approach. The posterior approach was also chosen to allow orthopaedists to work with a single access, while patients with particularly complicated situations, especially in posterior knee dislocations or pluriframmentary fractures, required the medial approach. In every surgery, systemic heparinization was performed, except in a haemophilic patient with increased risk of intraoperative bleeding. Treatments were performed according to the lesion, availability of the great saphenous vein (GSV) of adequate calibre (at least 3 mm in diameter), and, where possible, it was preferred to perform end-to-end anastomosis of the popliteal artery. Three patients were treated with GSV graft, four by end-to-end anastomosis of the popliteal artery without any graft, and three by synthetic grafting due to an inadequate GSV calibre. Revascularization of the popliteal artery was successful in seven cases out of 11, where two patients underwent amputation above the knee. One of the two patients was a 54-year old obese woman (BMI=40 kg/m2) who had rheumatoid arthritis and drug-induced osteoporosis due to corticosteroid therapy and who experienced low-velocity trauma from falling. Her knee prosthesis was dislocated after her fall from an upright position, which injured her popliteal artery. Because of her immune dysfunction due to rheumatoid arthritis and she was administered corticosteroid therapy, she was affected by a multi-drug resistant infection of the surgical wound that made amputation necessary after 2 months of medical therapy. A second patient was 24-year-old man who attempted suicide by falling from a height of 12 meters and who underwent bilateral GSV graft revascularization and osteoarticular stabilization but who ultimately required above-knee amputation due to extensive soft tissue injury. A third patient was worsened by haemophilia type A died 6 months after GSV procedure due to the extensive injuries caused by the accident. The septic shock despite antibiotic prolonged therapy, together with traumatic soft tissue injuries, resulted in the patient’s death, but graft patency was maintained. Cases of patients treated with revascularization due to a popliteal artery injury are described in Table 2, which shows that there was no difference in revascularization success between high-velocity and low-velocity trauma patients. All patients were intraoperative submitted to ecocolor doppler. Intraoperative angiography was used in only one patient. Two patients needed above-knee amputation before 30 days after the procedure: one due to infection of the surgical access and the other due to severe soft tissue injury. Revascularization of the popliteal artery was successful in seven cases out of 11, where two patients underwent amputation above the knee. A second patient was 24-year-old man who attempted suicide by falling from a height of 12 meters and who underwent bilateral GSV graft revascularization and osteoarticular stabilization but who ultimately required above-knee amputation due to extensive soft tissue injury. A third patient was worsened by haemophilia type A and died 6 months after GSV procedure due to the extensive injuries caused by the accident, as well as surgical wound infection. Despite the prolonged antibiotic therapy, septic shock together with traumatic soft tissue injuries, resulted in the patient’s death, but graft patency was maintained. Cases of patients treated with revascularization due to a popliteal artery injury are described in Table 2, which shows that there was no difference in revascularization success between high-velocity and low-velocity trauma patients. One patient died during hospitalization due to trauma-related complications and comorbidities, namely haemophilia type A, despite maintaining graft patency. Among the complications, there were two patients with deep venous thromboses (6.4%), and two patients (6.4%) healed by secondary intention. Follow-up was available for 6 of 8 salvaged limbs over a mean 12-month period where all patients maintained patency of arterial reconstruction, vein graft, or synthetic graft. Further, 1-year patency was maintained in three cases of end-to-end anastomoses, one case of GSV graft, and two cases of synthetic grafting.

**Table 2:** Patients with traumatic popliteal artery injury. \**Pt. n°8-9: bilateral lesion*.

| *Pt* | *sex* | *age* | **BMI**<br>**(Kg/m<sup>2</sup>)** | **Trauma**<br>**dynamic** | **Osteoarticular**<br>**Trauma** | **Revascularization** | **Postoperative**<br>**course** | **comorbidity** |
| --- | --- | --- | --- | --- | --- | --- | --- | --- |
| 1 | F | 54 | 40,2 | Low speed | Knee dislocation of the knee prosthesis | End-to-end | Amputation above knee | Reumatoid arthritis, osteoporosis, total knee prosthesis, hip prosthesis (bilateral) |
| 2 | M | 35 | 27 | High speed | Proximal tibial epiphysis fracture | GSV graft | Discharged. Maintaining patency. | — |
| 3 | M | 65 | 31,5 | High speed | Knee dislocation of the knee prosthesis | ePTFE synthetic graft | Discharged. Maintaining patency | Arthrosis, total knee prosthesis |
| 4 | M | 52 | 25 | High speed | Distal femoral epiphysis fracture, proximal tibial epiphysis | ePTFE synthetic graft | Discharged. Maintaining patency | — |
| 5 | M | 57 | 18,7 | High speed | Knee dislocation, vertebral L1 fracture, pelvis fracture | GSV graft | Deceased due to traumatic soft tissue lesions, infection and sepsis. | Haemophilia A |
| 6 | M | 22 | 26,6 | High speed | Proximal tibial epiphysis fract., tibial diaphysis frac., fibula fracture | End-to-end | Discharged. Maintaining patency | — |
| 7 | F | 57 | 35,8 | Low speed | Knee dislocation | GSV graft | Discharged. Maintaining patency |  |
| 8 | M* | 24 | 25 | High speed | Proximal tibial epiphysis fracture, Distal femoral epiphysis fracture, tibial multifragmentary fracture | GSV graft | Amputation above knee due to extensive soft tissue lesion. |  |
| 9 | M* | 24 | 25 | High speed | Distal femoral epiphysis fracture, tibial multifragmentary fracture | GSV graft | Discharged. Maintaining patency |  |
| 10 | F | 71 | 23 | High speed | Proximal tibial epiphysis fracture | End-to-end | Discharged. Maintaining patency | Arthrosis, osteoporosis |
| 11 | M | 37 | 28,3 | High speed | Colles fracture | ePTFE synthetic graft | — | — |

## Discussion

Popliteal artery traumas represent a minority of arterial traumas likely due to their anatomical position in the popliteal fossa and their location, which is posterior to the knee joint. Unless there are pre-existing structural lesions (aneurysms) or genetic predispositions such collagenopathy, Ehlers-Danlos Syndrome, etc… (laxity of joint ligaments), high-velocity traumas are required to disrupt the knee joint. Although rare, cases of low-velocity traumas exist; these can cause damage to the joint and involve the popliteal artery. This is dangerous because of the low velocity of trauma associated with a large weight can be equally dangerousand inconsistent signs and symptoms; typical signs of arterial injury are absent in up to 40% of patients, where the presence of a pulse in the affected limb does not exclude an arterial lesion and segmental Doppler may not detect the injury16–19. This type of injury does not depend only on the trauma dynamic (as with high-velocity traumas) but instead on the patient’s risk factors. According to the literature, the keys for effective revascularization are systemic heparinization20–22, performing revascularization between 6 and 8 hours9, and end-to-end anastomosis instead of using a GSV graft or synthetic graft. Wherever possible, we preferred end-to-end anastomosis over GSV grafting because it is associated with a higher success rate13. Furthermore, several authors have suggested that fibromuscular tethering of the artery in the popliteal fossa precludes tension-free anastomosis, unless potentially critical perigeniculate collaterals are divided. Therefore, they advocate interposition or bypass grafting in such cases20,23,24. Extension of soft tissue injuries is crucial for adequate and successful revascularization; this is the reason amputation rates due to reason arterial injury from secondary blunt trauma are double those due to injury from penetrating trauma25. Both patients affected by low-velocity trauma had a BMI greater than 35 kg/m2 while patients affected by high-velocity trauma had a BMI between 18.7 and 31.5 kg/m2. The advanced age of patients affected by low-velocity trauma could represent a predisposing factor with regard to diseases associated with aging such as osteoporosis, arthrosis, and ligamentous laxity, rendering them prone to fractures or dislocations, even of the knee joint, as body weight represents a strong risk factor for low-velocity trauma fractures and dislocations8,14,26. In three cases of our experience patients had a knee prosthesis dislocation, a rare but possible cause of popliteal artery injury, especially in posterior dislocations; with this condition the disruption of ligaments, is also described by Bonnevialle et coll. in concomitant palsy of the common peroneal nerve 27,28. Our findings align with the those in the literature13 in that there was no correlation between the dynamics of the trauma and the success of revascularization. This study has some limitations. The first was the small number of patients since our Institution is not a trauma centre and popliteal artery traumas have a low incidence. Furthermore, as in the literature reviews, there is no single definition of low/high-velocity trauma. Therefore, different articles may differ in results depending on their interpretation. Finally, as patients in this study were from a specific population, the findings about mass/velocity data cannot be extrapolated to the general population. The statistically significant conclusions will be drawn only after more data collection. However, this correlation remains very suggestive and can be deepened with further studies.

In conclusion, traumatic lesions of the popliteal artery following low-velocity trauma are correlated with high BMI of >35 kg/m2 and must considered in orthopaedic evaluation. Secondly, trauma dynamics (low- or high-velocity) do not influence the success of revascularization. Instead, factors that influence the results are revascularization procedure between 6 and 8 hours after the accident, intraoperative systemic heparinization, and an appropriate technique of revascularization and extension of soft tissue injuries. There may be an association between age and susceptibility to arterial trauma relative to low-velocity trauma; however many more risk factors should be analysed and a greater number of cases would be needed. Further in-depth studies should be conducted to support this association.

## Data Availability

Hospital electronic chart
Detailed operative electronic reports in Thaching hospital

## Author Contribution

AAM literature search, study design, data analysis, data interpretation, writing, critical revision

VN literature search data collection, writing

GC writing, critical revision

FDM data analysis, data interpretation, writing, critical revision

VP data analysis, data interpretation, writing, critical revision

PF, data interpretation, writing, critical revision

AI, data interpretation, writing, critical revision

All applicable section of the study was made following STROBE Statement regarding epidemiological studies

The authors declare no conflicts of interest.

The authors declare no funding was provided.

---

**Figure 1.**
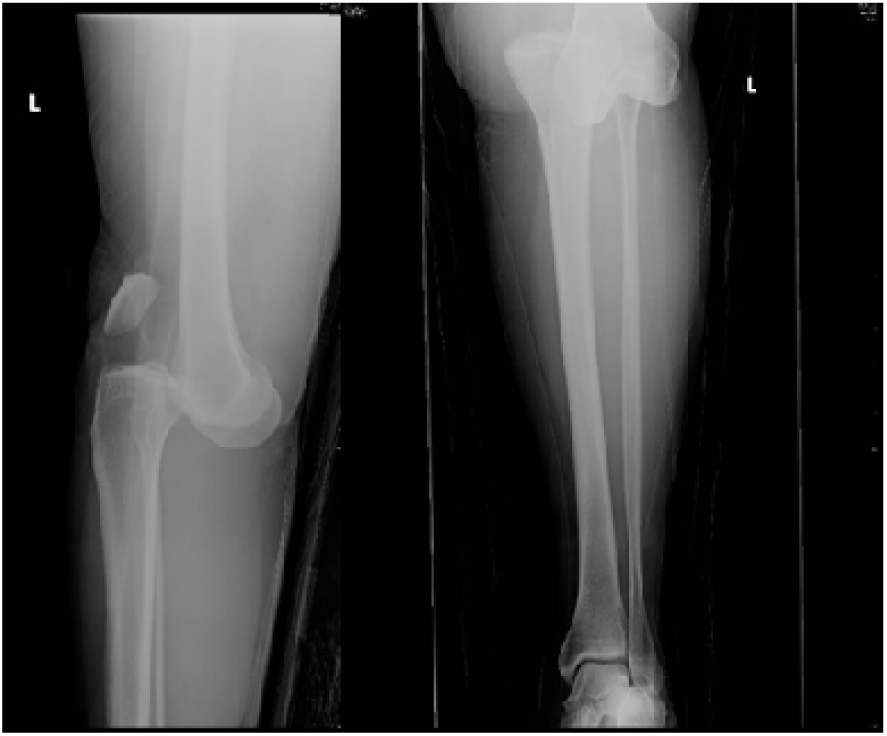
Pt n°7. Plain RX: Posterior dislocation of the knee. Note the posterior descent of the bone segment represented by the femur.

**Figure 2.**
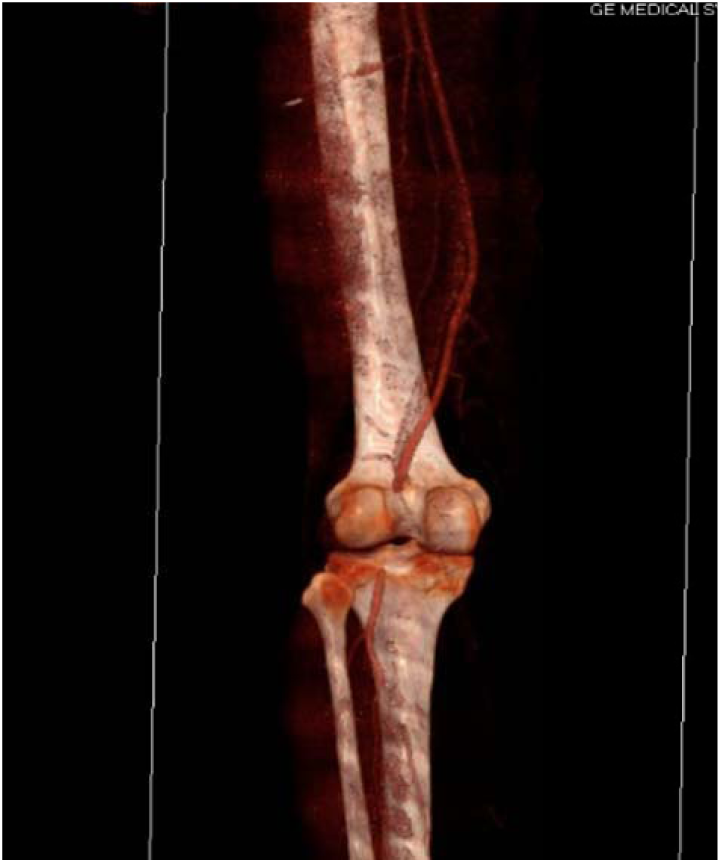
Pt n°7 CT scan of lower left limb: after reduction of dislocation, there is an interruption of the passage of the contrast medium in the retroarticular popliteal artery.

**Figure 3.**
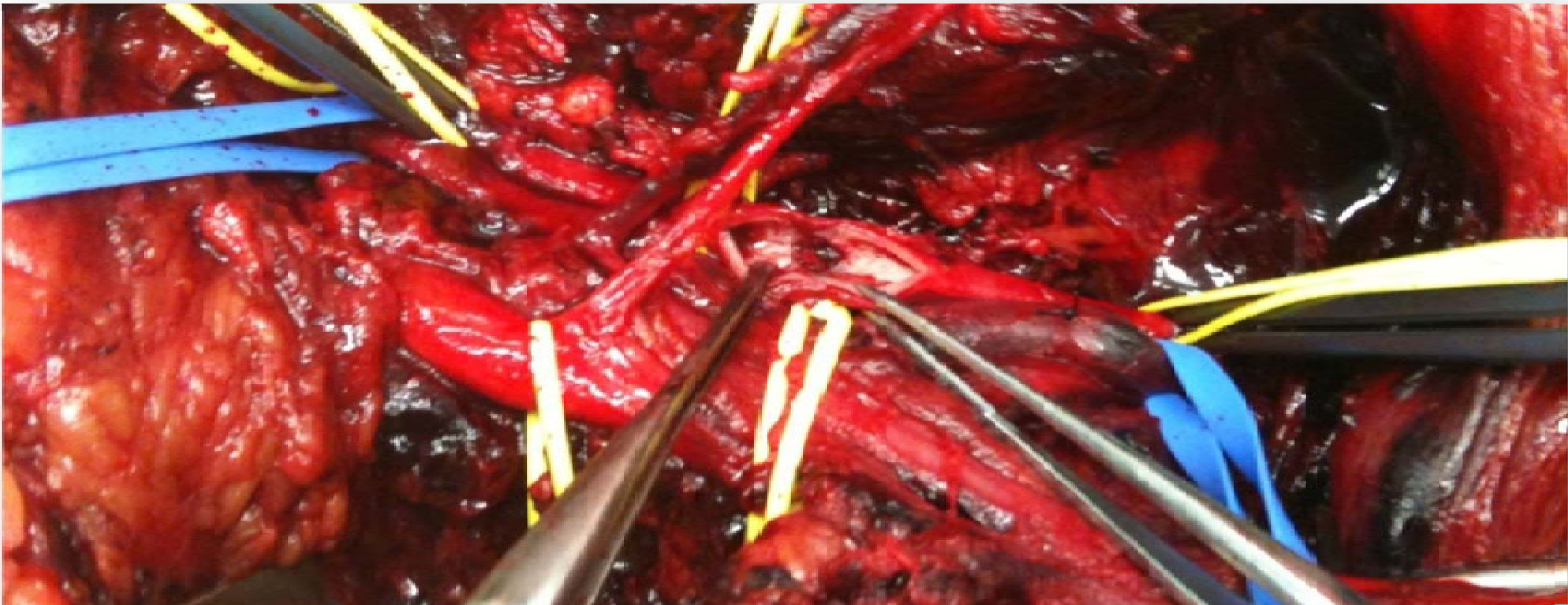
Pt n°7 Intraoperative image: posterior politeal surgical access. Popliteal artery clamping and longitudinal arteriotomy. After removal of the intraluminal clot, note the interruption of the tunica intima, the seat of indirect arterial trauma.

